# Vaccination and Non-Pharmaceutical Interventions: When can the UK relax about COVID-19?

**DOI:** 10.1101/2020.12.27.20248896

**Authors:** Sam Moore, Edward M Hill, Mike J Tildesley, Louise Dyson, Matt J Keeling

## Abstract

**Background:** The announcement of efficacious vaccine candidates against SARS-CoV-2 has been met with worldwide acclaim and relief. Many countries already have detailed plans for vaccine targeting based on minimising severe illness, death and healthcare burdens. Normally, relatively simple relationships between epidemiological parameters, vaccine efficacy and vaccine uptake predict the success of any immunisation programme. However, the dynamics of vaccination against SARS-CoV-2 is made more complex by age-dependent factors, changing levels of infection and the potential relaxation of non-pharmaceutical interventions (NPIs) as the perceived risk declines.

**Methods:** In this study we use an age-structured mathematical model, matched to a range of epidemiological data in the UK, that also captures the roll-out of a two-dose vaccination programme targeted at specific age groups.

**Findings:** We consider the interaction between the UK vaccination programme and future relaxation (or removal) of NPIs. Our predictions highlight the population-level risks of early relaxation leading to a pronounced wave of infection, hospital admissions and deaths. Only vaccines that offer high infection-blocking efficacy with high uptake in the general population allow relaxation of NPIs without a huge surge in deaths.

**Interpretation:** While the novel vaccines against SARS-CoV-2 offer a potential exit strategy for this outbreak, this is highly contingent on the infection-blocking (or transmission-blocking) action of the vaccine and the population uptake, both of which need to be carefully monitored as vaccine programmes are rolled out in the UK and other countries.

**Research in context:** *Evidence before this study:* Vaccination has been seen as a key tool in the fight against SARS-CoV-2. The vaccines already developed represent a major technological achievement and have been shown to generate significant immune responses, as well as offering considerable protection against disease. However, to date there is limited information on the degree of infection-blocking these vaccines are likely to induce. Mathematical models have already successfully been used to consider age- and risk-structured targeting of vaccination, highlighting the importance of prioritising older and high-risk individuals.

*Added value of this study:* Translating current knowledge and uncertainty of vaccine behaviour into meaningful public health messages requires models that fully capture the within-country epidemiology as well as the complex roll-out of a two-dose vaccination programme. We show that under reasonable assumptions for vaccine efficacy and uptake the UK is unlikely to reach herd immunity, which means that non-pharmaceutical interventions cannot be released without generating substantial waves of infection.

*Implications of all the available evidence:* Vaccination is likely to provide substantial individual protection to those receiving two doses, but the degree of protection to the wider population is still uncertain. While substantial immunisation of the most vulnerable groups will allow for some relaxation of controls, this must be done gradually to prevent large scale public health consequences.

## 1 Introduction

The outbreak of SARS-CoV-2 that began in Wuhan, China, in late 2019 dramatically shaped life in 2020 as a world-wide pandemic emerged ^1^. In the UK, the first cases were identified on 31st January 2020^2^, with February and March witnessing an exponential rise in cases ^3^. The first lockdown began on 23rd March and reversed the growth in infection, although important health metrics such as hospital occupancy and deaths continued to increase for several days ^4^. The steady, but spatially heterogeneous, decline continued until August 2020 when a relaxation of controls and increased mixing as a result of this precipitated a second wave and, subsequently, a second lockdown in November 2020. By early December 2020, the UK had suffered over 60,000 deaths and 225,000 hospital admissions owing to COVID-19 and yet it is estimated that less than 20% of the population has been exposed to the virus ^5^, suggesting the outbreak is far from complete. Mass vaccination, and hence protection, of the population offers a potentially rapid exit strategy.

Within a year, over 50 companies have developed the first vaccines against any coronavirus. In early December 2020, one of these (Pfizer/BioNTech’s BNT162b2^6^) was approved for use in the UK, with several others in late Phase 3 trials at that time showing promising preliminary efficacy data. As of 12th December 2020, the UK had ordered 357 million doses of vaccines from 7 different developers: 100 million doses of University of Oxford/AstraZeneca vaccine ^7^; 40 million doses of BioNTech/Pfizer vaccine; 7 million doses of Moderna vaccine; 60 million doses of Novavax vaccine; 60 million doses of Valneva vaccine; 60 million doses of GSK/Sanofi Pasteur vaccine; and 30 million doses of Janssen vaccine ^8^. This is far in excess of any possible demand from the UK population, but mitigates for potential delays or failures from any single manufacturer. One continuing unknown with the potential vaccines is the degree to which they impact onward transmission (rather than simply preventing symptomatic infection); this is a key uncertainty that is investigated throughout this paper.

Vaccination against SARS-CoV-2 provides multiple unique challenges that are not encountered by other vaccination programmes. Most of the intuition about vaccination programmes is based on childhood vaccines where the aim is simply to achieve high uptake in each birth cohort and associated boosters. To date the seasonal influenza programme represented the largest annual delivery of vaccination in the UK ^9^, but seasonal influenza immunisation is pro-active (beginning before many cases arise), the ‘flu’ season is of relatively short duration and *R* for seasonal influenza is lower than for SARS-CoV-2 owing to both greater population immunity and a lower basic reproduction ratio ^10^. In contrast, for SARS-CoV-2 there is a race between infection and vaccination - vaccination rates are limited by supply and logistics, whereas infection can grow exponentially. However, the infection rate can be reduced by a range of non-pharmaceutical interventions (NPIs) whilst a vaccine can be targeted to where it will have the most impact ^11^. The future of COVID-19 control is therefore dependent, in a complex non-linear way, on the initial prevalence of infection, the level of NPIs and therefore the rate of growth or decay, the speed with which the vaccine can be rolled out, the targeting and uptake of the vaccine and the vaccine characteristics. The uncertainties and interactions between these components necessitates the use of mathematical models to explore scenarios.

Here, we present an age-structured mathematical model, matched to a range of epidemiological data, to forecast the dynamics of COVID-19 in 2021 and beyond based on multiple combinations of scenarios. These model results provide likely bounds on the expected number of deaths and hospitalisations, thus providing important policy insights into the interaction between continued non-pharmaceutical interventions and the ongoing vaccination programme. We focus on the risk-structured delivery programme for the UK and the potential risks of relaxing NPIs; we also consider the individual risks and how these are mitigated by vaccination.

## 2 The mathematical model and vaccination assumptions

We adapt an existing age and regionally structured model of SARS-COV-2 dynamics that has been matched to UK data ^12,13^ to include the consequences of vaccination ^11^. The model captures the historic trends of infection, hospitalisation and deaths in the UK, including the scale of the first and second waves. Including vaccination into this model shows how prioritising the oldest age-groups leads to the greatest reduction in deaths ^11^. Here we increase the realism of the vaccination dynamics, including the timing of vaccine roll out across the population and the need to administer two doses. One key issue that the model cannot address is the level of NPIs that will be imposed in the future and the level of support (and therefore adherence to NPIs) from the general population. We have optimistically assumed that controls in the short term are sufficient to keep the reproductive number (*R*) just below one; we then relax controls at various times throughout 2021 to investigate the level of protection generated by vaccination. All results represent the mean of multiple simulations which explore the inferred epidemiological parameter space determined by matching to a range of epidemiological time series; credible intervals for the predictions are shown in the Supplementary Material.

Vaccination schedules for the UK are still not determined over long time scales, although the immediate priority order has been defined ^11,14^. We implement an accelerating delivery programme that approximates plausible roll-out of SARS-CoV-2 vaccination in the UK (Figure 1(a)), following in each stage the identified priority ordering:

**Figure 1:**
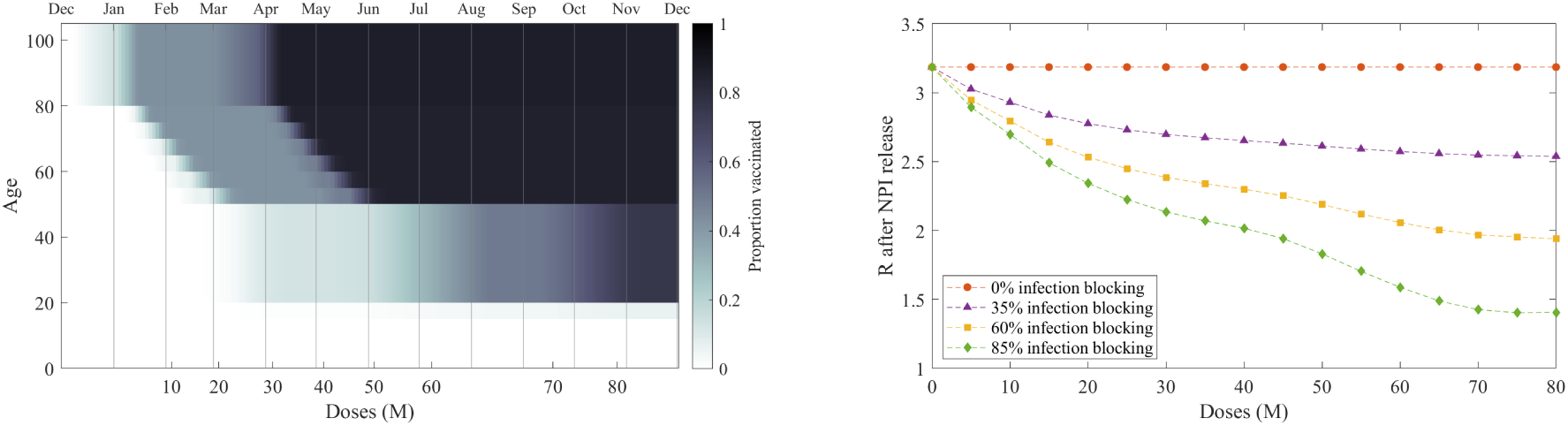
Scheduling and impact of vaccine uptake. **(Left)** The assumed uptake over time showing the proportion in each age-group that have received two doses of vaccine by the total number of doses (lower x-axis) or a given date (upper x-axis). Darker shading corresponds to a greater proportion vaccinated. **(Right)** The impact of the vaccination programme, defined by the number of administered doses, on the reproductive ratio *R* in the absence of all non-pharmaceutical interventions. We display four different assumptions about the ability of the vaccine to block infection and hence stop further transmission: 0%, 35%, 60% and 85% reduction in infection due to two doses of the vaccine.

- 1 million doses of Pfizer/BioNTech vaccine alone across December.
- 1 million doses per week from the start of January, increasing to 2m per week by February using a mixture of vaccines.
- 2 million doses per week from the start of February until vaccine completion using a mixture of vaccines.

Throughout, we assume 95% uptake in care homes, 85% elsewhere above the age of 50 and 75% below the age of 50 for dose 1, dropping to 75% and 66% for above and below 50 respectively for dose 2. In practice, vaccination is also likely to be highly correlated within households and socio-demographic groups ^15^, which will weaken the population-scale impact of any infection blocking by the vaccine.

We use a 2-dose model to simulate the impact of vaccination in both reducing infection (and hence onward transmission) and in reducing symptomatic disease. We assume that delivery of the second dose is prioritised over new first doses, with a minimum 84 day delay between doses (Figure 1(a)). In the absence of detailed vaccine specific information, we also assume a stepped efficacy over time following the first dose, which scales with the assumed final vaccine efficacy (VE): from the first dose to day 14, zero efficacy; from day 14 to second dose on day 84, 80% of VE; from day 84 to day 98, 80% of VE; from day 98 onwards, 100% of VE (Supplementary Material).

Vaccine efficacy against disease is assumed to be high (in keeping with preliminary reports): 94% during the earliest phase where just the Pfizer/BioNTech vaccine is used, dropping to 89% (on average) when multiple vaccines are in use. The role of vaccines in blocking infection, and hence onward transmission, is less clear so we consider a range of infection efficacy from 0% to 85%, which we assume operates by preventing primary infection. We note that the disease efficacy takes into account both infection blocking and the reduction of severe symptoms if infection does occur (Supplementary Material).

While efficacy against disease is of immediate benefit in protecting individuals against developing severe symptoms, it is the infection blocking potential of the vaccine that leads to a reduction in the intrinsic growth rate and the reproduction number, *R*. Figure 1(b) shows the reproduction number *R*, on release of all non-pharmaceutical interventions, as the vaccine programme progresses (but ignoring any additional increase in immunity from infection). Four different assumptions for infection efficacy (infection blocking) are considered: for zero infection efficacy (orange line), *R* ≈ 3.2, this is higher than in the first wave due to the new variant but is reduced from its theoretical maximum due to the slight depletion in susceptibles from natural infection up to January 2020; whereas when infection efficacy is high (85% infection blocking, green line), vaccination can generate a substantial decline in the reproductive number, although still insufficient to drive *R* below 1 for our default assumptions about vaccine uptake.

## 3 Predictions under vaccination and changing NPIs

We simulate the infection dynamics from February 2020, matching to the observed pattern of cases, hospitalisations and deaths until the middle of January 2021, and then predict the impact of vaccination on daily deaths until the end of 2021 (Figure 2). Unsurprisingly, subsequent waves of infection and associated deaths are reduced by increasing levels of infection efficacy. Early, modest relaxation of NPIs (Figure 2(a)), matched to the levels observed in early September (when *R* was between 1.2 and 1.4 across different English regions and devolved nations with the old variant in predominant circulation) leads to subsequent waves of infection even under the most efficacious assumptions (as displayed by the green trace, corresponding to a vaccine that blocks 85% of infection). Later relaxation of NPIs (April 2021 in Figure 2(b)) provides a greater opportunity for some level of herd-immunity to have accrued if the vaccine is moderately effective at blocking infection.

**Figure 2:**
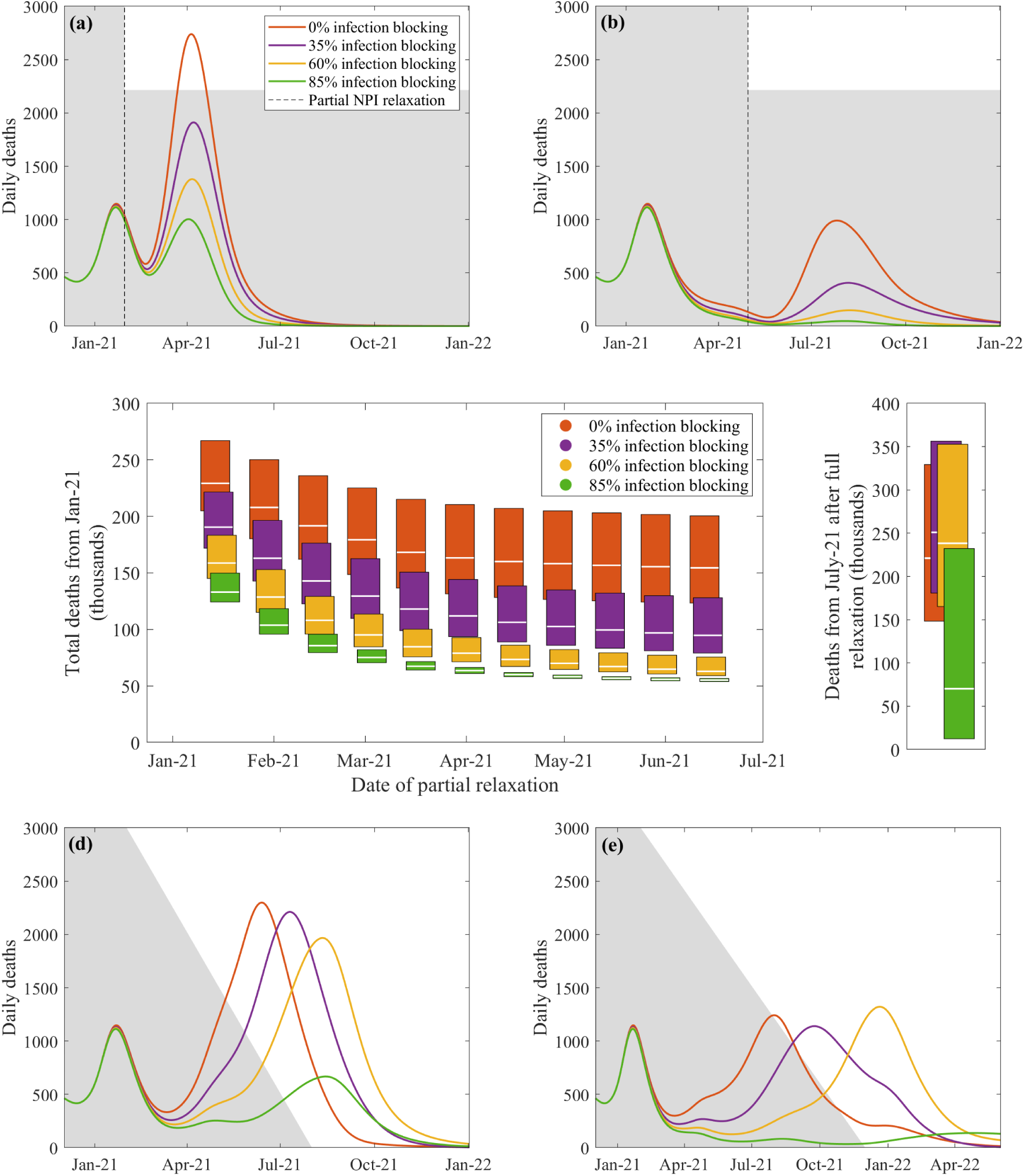
Predicted daily deaths in the UK following the start of an immunisation program and relaxation or removal of NPIs. Panels **(a-b)** show the effect of relaxing current NPI measures down to those seen in early September 2020 (generating *R*∼ 1.2 − 1.6 dependent upon the region and level of the new variant) from February-May 2021 respectively. Panel **(c)** displays the aggregate effects of partial release of NPI measures at different dates during the vaccination programme (left) compared with complete release from July 2021 (right); the upper limit, central bar and lower limit of each box corresponds to pessimistic, default and optimistic assumptions about vaccine uptake. The lower panels **(d-e)** correspond to a gradual reduction in NPIs until removal of all controls, as illustrated by the grey area. The default scenario (panels **(a**,**b**,**d**,**e)**) assumed 85% and 75% uptake in those above and below 50 years respectively; the optimistic scenario increased this uptake to 95% and 80% in the two age-groups; while the pessimistic scenario decreased this to 70% and 60% uptake respectively.

To consider sensitivity to vaccine uptake, we analyse the total number of deaths predicted by the model (Figure 2(c)), which equates to the areas under the curve in the preceding graphs. The central bar represents 85%/75% and 75%/66% uptake above and below the age of 50 for dose 1/2 respectively (as shown in Figure 1 and the rest of Figure 2). The lower limits of each box correspond to more optimistic uptake, corresponding to an increased 95% and 80% first dose uptake above and below 50 years old respectively, whilst the upper limits represent a more pessimistic scenario with 70%*/*60% dose 1 uptake. In both additional scenarios we retain the relative 12% decrease in uptake for dose 2, in line with the central scenario.

Assessing the predicted mortality from the start of 2021 for different dates at which NPI are partially relaxed (to the level observed in early September 2020), even maintaining these levels of NPI control and having a highly efficacious vaccine we estimate over fifty thousand deaths are likely to occur from January 2021 due to the slow decline in cases from its current high level (Figure 2(c), left-hand panel); early relaxation of control measures or low infection efficacy can lead to a pronounced subsequent wave of infection.

If we wish to completely lift *all* restrictions once both phases of the vaccination campaign are complete, we predict a substantial outbreak with a large number of associated deaths (Figure 2(c), right-hand panel). When the vaccine is not infection blocking, removing NPIs triggers an uncontrolled wave of infection in which only those successfully immunised (approximately 88% of 85%) will escape. Optimistic assumptions for vaccine efficacy may still lead to a further 123 thousand deaths (54 thousand from January to July, and 69 thousand from July onwards when NPIs are released) at our expected uptake level.

The stepwise release of all NPIs (Figure 2(c) right-hand panel, Supplementary Material) modelled so far generates an over-shooting wave of infection; a more gradual release of restrictions mitigates these effects (Figure 2(d-e)). A slow release of NPIs (as illustrated in Figure 2(e)) generates the fewest deaths, with around 99 thousand more deaths (from January 2021) forecast under our most optimistic efficacy assumptions.

The precise dynamics and outcomes are contingent on the assumed intrinsic growth rate before the relaxation of NPIs, which here is approximately *R* = 0.8 following a tight January lockdown — other values for this quantity will change the precise curves but would not change the qualitative conclusions. We see similar behaviour if we examine the number of daily hospitalisations (Supplementary Material), with a notable third wave predicted if NPIs are relaxed too early or if the vaccine has a limited impact on infection and hence onward transmission.

## 4 Impact of vaccination status

Whilst we have predominantly focused on the population-level consequences of the vaccination programme, one key question is the likely vaccination status of individuals that are severely ill. We characterise this relationship as a function of the number of doses delivered so far (as part of the mass vaccination programme) and for 85%/75% and 75%/66% uptake above and below the age of 50 for dose 1/2 respectively (Figure 3). We display findings assuming 50% infection efficacy, with similar results obtained for all levels of infection efficacy. We consider four categories of individual: those who have not yet been offered the vaccine; those who are in an eligible age-group but due to health reasons or personal beliefs remain unvaccinated; those who had received one dose so far; and those that have received both doses. Between 0-30M doses and 55-70M doses those individuals that have not been offered the vaccine declines linearly, while those unvaccinated but in eligible age groups and those that have received two doses grow linearly. In the interim and later periods first doses are instead replaced by second doses (Figure 3(a)).

**Figure 3:**
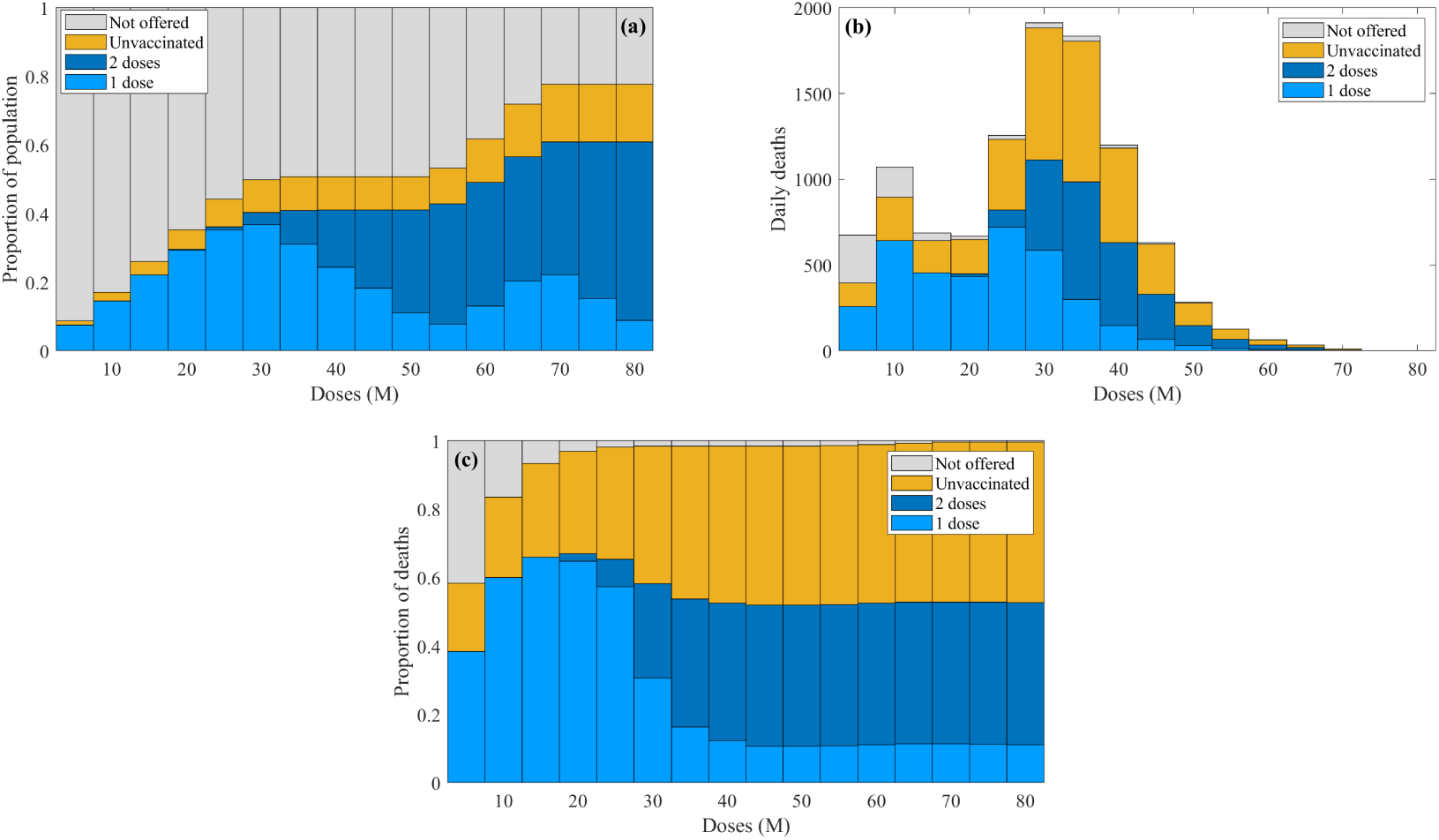
Characterisation of the dynamics in terms of vaccine status as a function of the number of doses delivered so far. We consider four categories of individual: those who have not yet been offered the vaccine (grey); those who are in an eligible age-group but due to health reasons or personal beliefs remain unvaccinated (yellow); those who had received one dose so far (light blue); and those that have received both doses (dark blue). The composition of the entire population; **(b)** The number of daily deaths; **(c)** The proportion of deaths. We display simulations assuming 85%/75% and 75%/66% uptake above and below the age of 50 for the first and second vaccine dose respectively, 60% infection efficacy, and with a moderate reduction in NPIs at the start of Feb 2021 (corresponding to the yellow curve in Figure 2(a)).

The unfolding epidemic (matched to Figure 2(a)) is then distributed across these four status groups (Figure 3(b)). Owing to the fact that the existing vaccination strategy targets the most vulnerable first, deaths are very quickly dominated by those who have either received the first vaccine dose or those who have not been vaccinated but are in eligible groups, with those who have not been offered the vaccine accounting for less than 10% of deaths by the time that 15M doses have been administered.

By plotting the proportion of all deaths associated with each status we observe that around 40% of deaths can be expected in those that have been vaccinated (Figure 3(c)). We stress that while at the individual-level two doses of vaccine reduces the risk of mortality by 80%, because vulnerable vaccinated people rapidly outnumber vulnerable unvaccinated people we should expect to see a high proportion of vaccinated individuals suffering severe disease and mortality.

The final shape of these distributions is a function of vaccine uptake in the most at-risk; increasing vaccine uptake reduces the number of deaths but paradoxically increases the contribution of vaccinated individuals to the proportion of deaths. There is also a strong influence of how well the vaccine protects against severe disease – greater efficacy against the most severe disease will again reduce the number of deaths and will also decrease the proportions associated with vaccinated individuals. However, if the vaccine is less efficacious in the elderly this trend would be reversed.

## 5 Conclusions

Here we have shown that high efficacy vaccines that provide a substantial level of infection blocking offer a means of eventually relaxing controls without suffering a large subsequent wave of hospitalisations and deaths. Our conclusions rely on not only the vaccine characteristics but also upon the uptake in the population (Figure 2(c)); in particular the most vulnerable groups who require protection against disease, but also in the general population if infection blocking is to be successful. In practice vaccine uptake is likely to be regionally and socio-demographically correlated ^16,17^. Such correlations may potentially lead to pockets of high susceptibility in the population which can act as locations of small-scale outbreaks and reduce herd immunity ^18^. It is also likely that vaccine uptake will vary in time as the population’s perceived risk varies ^19,20^, with high levels of hospitalisations and deaths leading to a greater demand for the vaccine. We expect there to be a complex four-way interaction between levels of infection, NPI policy, NPI adherence and vaccine uptake. From a public health perspective, it is therefore key to understand the drivers of vaccine uptake and vaccine hesitancy, identify groups that may have lower than average uptake and plan accordingly.

Early relaxation of non-pharmaceutical interventions (NPIs), before sufficient immunity has been established is shown to precipitate a large wave of infection with resultant hospital admission and deaths; a similar impact is predicted from any final release of NPIs if herd immunity has not been achieved (Figure 2). Even with high levels of vaccine uptake, a very large fraction of the population needs to be immunised to prevent subsequent waves of infection (Figures 1 and 2), implying strong NPIs would still be required even when Phase 1 of the vaccination programme is complete to avoid surges in infection. A more measured approach in which NPIs are gradually released over a period of many months has advantages over sudden changes to controls, but may not mitigate the worst effects (Figure 2 and Supplementary Material). We stress that if hospital occupancy and deaths increased due to changes in NPIs, we would expect both national legislation and emergent behaviour to limit the spread ^21^, therefore our scenarios represent a pessimistic view of measures in response to a worsening outbreak.

As of January 2021, multiple vaccine manufacturers now have peer-reviewed publications presenting the findings of their phase 3 trials ^6,7,22^. These have been used to provide approximate parameters for this model-based study, but many questions have not been quantitatively addressed. A number of key vaccine parameters within the model are therefore based on parsimonious assumptions, and we identify the following three issues that require additional experimental data to refine model assumptions. Firstly, as elucidated throughout this paper determining whether the vaccine blocks infection is key for the development of herd immunity and hence the role of vaccination in the long-term control of COVID-19. There is also the potential for the vaccine to further reduce viral shedding from vaccinated individuals, hence reducing onward transmission, but this is likely to be difficult to measure. Secondly, we have assumed that efficacy against disease applies equally across the entire spectrum of disease, however if the vaccine has differential protection against the most severe disease this will impact our predictions for hospital admissions and deaths. Finally, we expect efficacy to vary with age and between risk groups; incorporating such heterogeneity into models is key for more robust predictions.

Over longer time scales the possibilities of waning immunity and mutation may influence these predictions. Waning immunity, either naturally derived or from vaccination, may necessitate seasonal vaccination programmes against SARS-CoV-2 protecting the most vulnerable in a similar manner to seasonal influenza vaccines ^23^. We are again lacking in our fundamental understanding of SARS-CoV-2 epidemiology, in particular whether subsequent infections have the same severity as primary infections, as well as quantitative estimates of the duration of protection. Both of these elements can be factored into the prediction mechanisms, but without detailed evidence such long-term forecasts are speculation.

Effective vaccines with high uptake are likely to be an essential element in the long-term control and potential elimination of COVID-19. However, experience with other diseases has illustrated that elimination is difficult and generally requires a targeted multi-strategy approach ^24^. The same is likely to be true for SARS-CoV-2. While mass vaccination will inevitably reduce the reproductive number *R* and reduce disease prevalence, other measures, such as intensive test-trace-and-isolate, will be needed to target pockets of infection. Ultimately, whether we achieve eradication of SARS-CoV-2 is likely to depend on the long-term natural history of infection and the public health importance attached to this goal.

## Data Availability

The data were supplied from the CHESS database after anonymisation under strict data protection protocols agreed between the University of Warwick and Public Health England.

## Acknowledgements

This research was funded by the National Institute for Health Research (NIHR) [Policy Research Programme, Mathematical & Economic Modelling for Vaccination and Immunisation Evaluation, and Emergency Response; NIHR200411], the Medical Research Council through the COVID-19 Rapid Response Rolling Call [grant number MR/V009761/1] and through the JUNIPER modelling consortium [grant number MR/V038613/1]. The funders had no role in study design, data collection and analysis, decision to publish, or preparation of the manuscript.

The data were supplied from the CHESS database after anonymisation under strict data protection protocols agreed between the University of Warwick and Public Health England. The ethics of the use of these data for these purposes was agreed by Public Health England with the Government’s SPI-M(O) / SAGE committees.

All authors declare that they have no competing interests.

## Author contributions

**Conceptualisation:** Matt J. Keeling.

**Data curation:** Matt J. Keeling; Edward M. Hill.

**Formal analysis:** Sam Moore.

**Investigation:** Sam Moore.

**Methodology:** Sam Moore; Matt J. Keeling.

**Software:** Sam Moore; Matt J. Keeling.

**Validation:** Sam Moore; Matt J. Keeling; Edward M. Hill; Louise Dyson; Michael J. Tildesley.

**Visualisation:** Sam Moore.

**Writing - original draft:** Matt J Keeling.

**Writing - review & editing:** Sam Moore; Matt J. Keeling; Edward M. Hill; Louise Dyson; Michael J. Tildesley.

## SUPPLEMENTARY MATERIAL

### S1 The Mathematical Model

Here we present the basic model formulation that underpins the age-structured predictions of COVID-19 dynamics in the UK (Figure S1), and how the parameters of this model have been inferred from the available data. We used a compartmental age-structured model, developed to simulate the spread of SARS-CoV-2 within ten regions of the UK (seven regions in England: East of England, London, Midlands, North East & Yorkshire, North West, South East and South West; and the devolved nations: Northern Ireland, Scotland and Wales) [1], with parameters inferred to generate a good match to deaths, hospitalisations, hospital occupancy and serological testing [2]. The model population is stratified by age, with force of infection determined by the use of an age-dependent (who acquires infection from whom) social contact matrix for the UK [3, 4]. Additionally, we allow susceptibility and the probabilities of becoming symptomatic, being hospitalised and the risk of dying to be age dependent; these are matched to UK outbreak data. Finally, we account for the role of household isolation, by separating primary and secondary infections within a household (more details may be found in [1]). This allows us to capture household isolation by preventing secondary infections from playing a further role in onward transmission. Model parameters were inferred on a regional basis using regional time series of recorded daily hospitalisation numbers, hospital bed occupancy, ICU occupancy and daily deaths [2].

**Figure S1:**
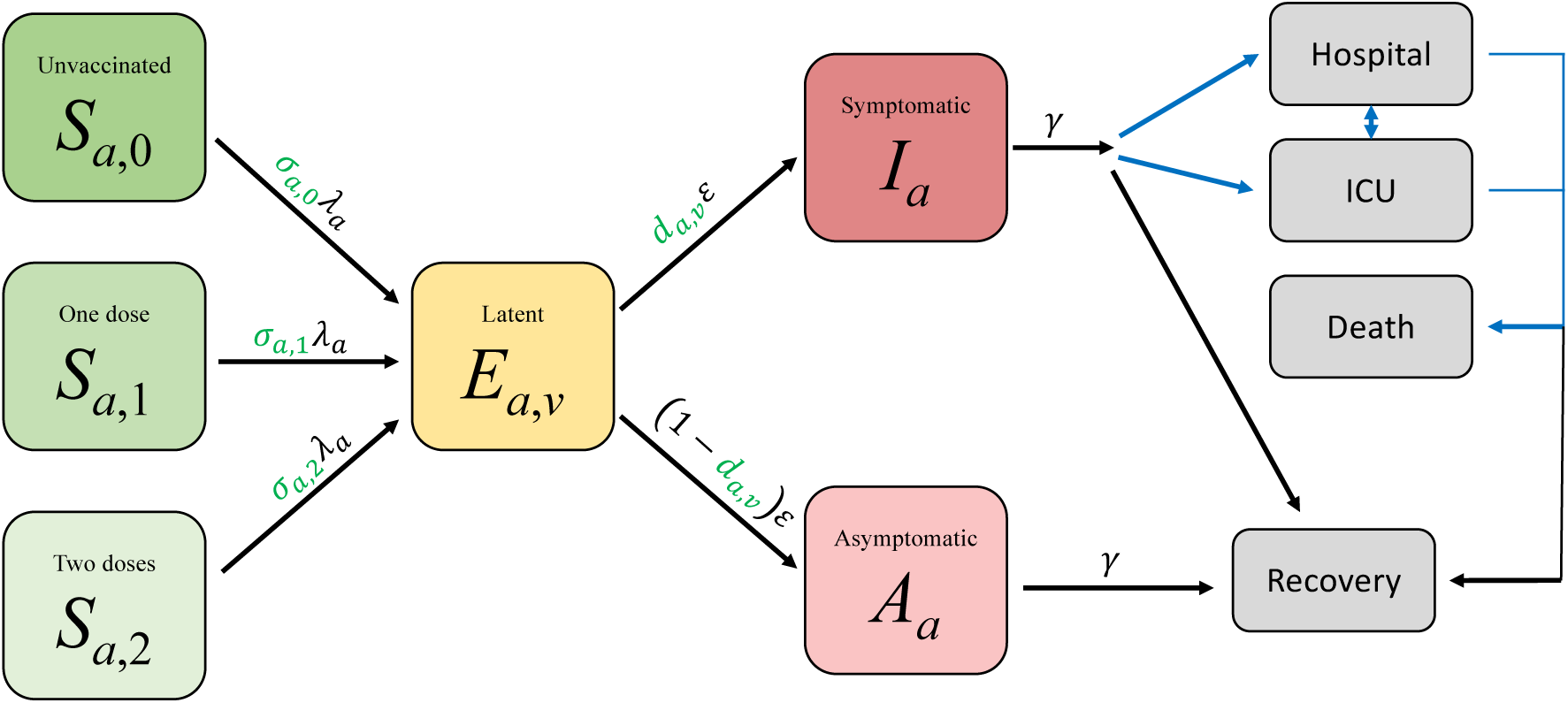
Representation of the basic model states and transitions. Black arrows show key epidemiological transitions while blue arrows show movements to observable states. Parameters in green show the action of vaccine on infection and probability of disease.

#### S1.1 Model description

We first show the underlying system of equations that account for the transmission dynamics, including symptomatic and asymptomatic transmission, household saturation of transmission and household quarantining. The population is stratified into multiple compartments: individuals may be susceptible (*S*), exposed (*E*), infectious with symptoms (*I*), or infectious and either asymptomatic or with very mild symptoms (*A*). Asymptomatic infections are assumed to transmit infection at a reduced rate given by *τ*. To some extent, the separation into symptomatic (*I*) and asymptomatic (*A*) within the model is somewhat artificial as there are a wide spectrum of symptom severities that can be experienced.

We let superscripts denote the first infection in a household (*F*), a subsequent infection from a symptomatic household member (*SI*) and a subsequent infection from an asymptomatic household member (*SA*). A fraction (*H*) of the first detected cases (necessarily symptomatic) in a household are quarantined (*QF*), as are all their subsequent household infections (*QS*) - we ignore the impact of household quarantining on the susceptible population as the number in quarantine is assumed small compared with the rest of the population. The recovered class is not explicitly modelled, although it may become important once we have a better understanding of the duration of immunity. We omitted natural demography and disease-induced mortality in the formulation of the epidemiological dynamics.

We extended the model formulation to capture a range of vaccination scenarios. We modelled two vaccination classes for individuals where it has been 14 days since they received their first and second dose of the vaccine; the 14-day delay allows partial immunity to develop (Figure S2). We included these within the *S* and *E* class by adding an additional vaccination subscript for the number of doses received; hence *S*_*a*,0_ corresponds to susceptible unvaccinated individuals while *S*_*a*,2_ corresponds to those that received their second dose of vaccine at least 14 days ago.

**Figure S2:**
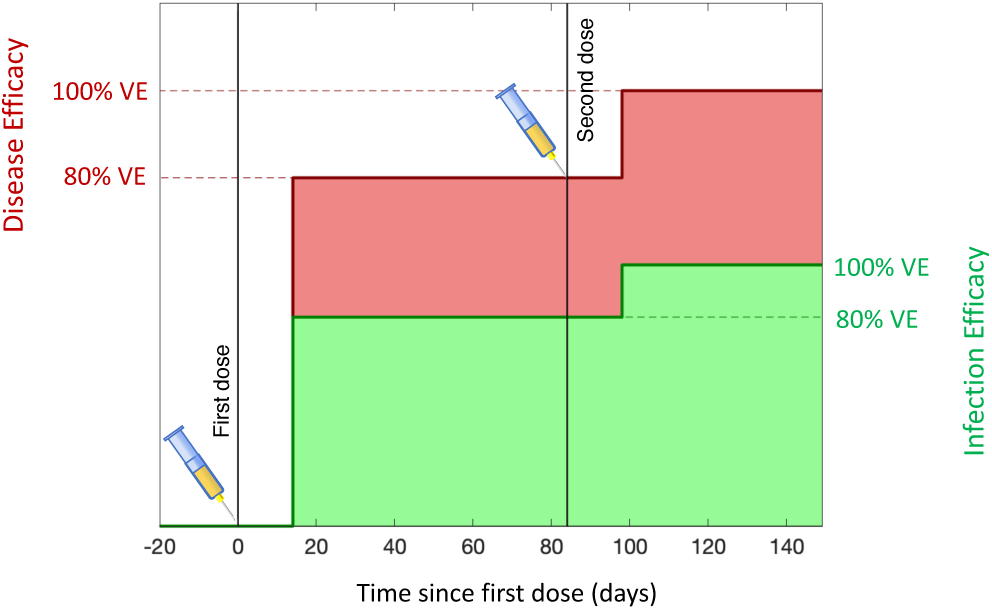
Dynamics of vaccine efficacy within an individual. 14 days after the first dose partial efficacy is developed, and 14 days after the second dose efficacy is raised to its maximum value. We highlight two forms of efficacy: disease efficacy (red) which prevents the development of symptomatic infection and acts on parameter *d* within the model; and infection efficacy (green) which prevents all infection and acts on parameter σ.

The full equations are given by

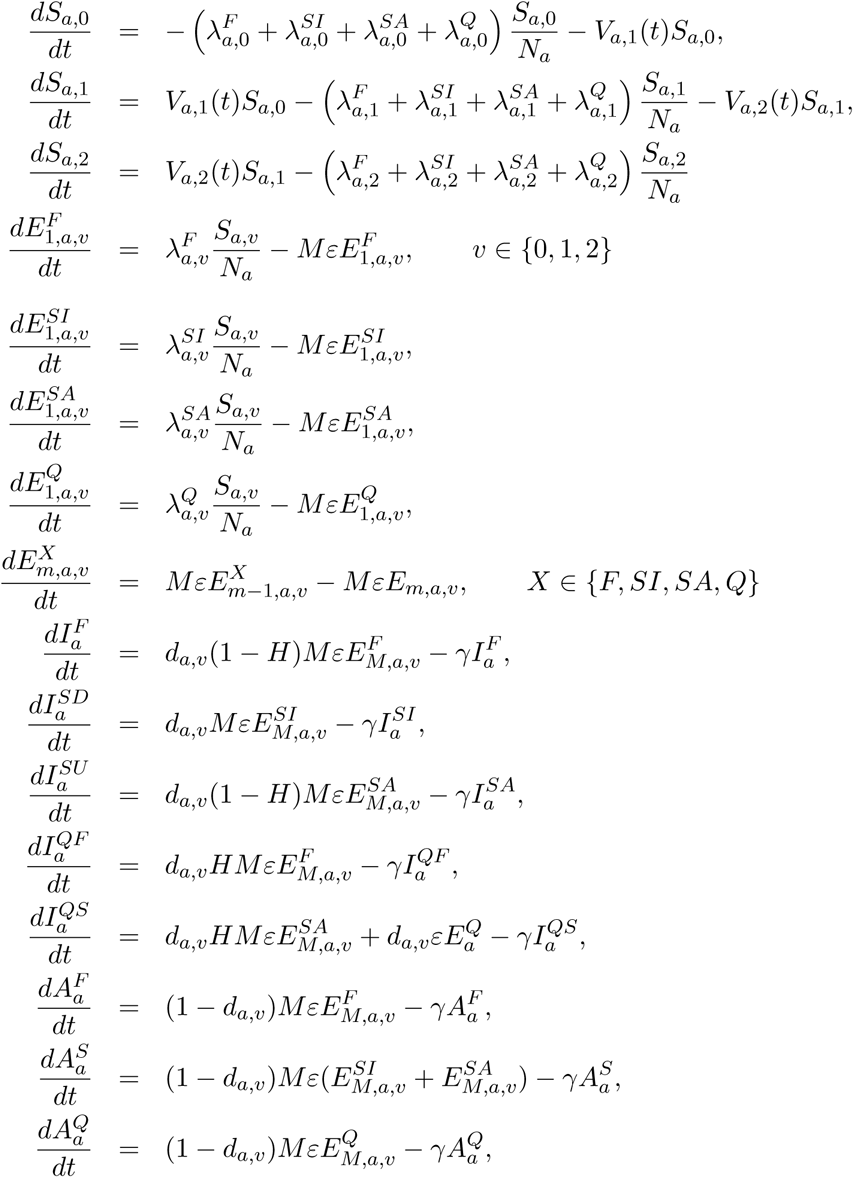

Here we have included *M* latent classes, giving rise to an Erlang distribution for the latent period, while the infectious period was exponentially distributed. Throughout we have taken *M* = 3.

The forces of infection which govern the non-linear transmission of infection obey:

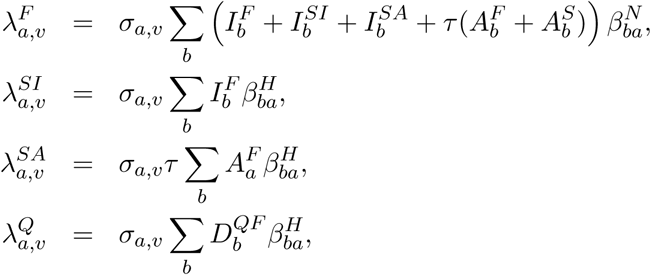

where *β*^*H*^ represents household transmission and *β*^*N*^ = *β*^*S*^ + *β*^*W*^ + *β*^*O*^ represents all other transmission locations, comprising school-based transmission (*β*^*S*^), work-place transmission (*β*^*W*^) and transmission in all other locations (*β*^*O*^). These matrices are taken from Prem *et al*. [4] to allow easily translation to other geographic settings, although other sources such as POLYMOD [3] could be used.

Two key parameters, together with the transmission matrix, govern the age-structured dynamics: σ_*a*_ corresponds to the age-dependent susceptibility of individuals to infection; *d*_*a*_ the age-dependent probability of displaying symptoms (and hence potentially progressing to more severe disease). Both of these are also modified by the vaccine status, such that those that have received one or two doses of vaccine have a lower risk of infection and a lower risk of developing symptoms. The action of vaccine on the parameter σ captures the infection blocking aspect of the vaccine, while the action on *d* captures the efficacy against disease (Figure S2). We also define *τ* as the reduced transmission from asymptomatic infections compared to symptomatic infections; given the probability of displaying symptoms is less in the younger age groups, this parameter shapes the role of younger ages in onward transmission.

We require our model to capture both individual level quarantining of infected individuals and isolation of households containing identified cases. In a standard ODE framework this level of household structure is only achievable at large computational expense [5, 6]. Thus, we instead made a relatively parsimonious approximation to achieve a comparable effect.

We assume that all within household transmission originates from the first infected individual within the household (denoted with a superscript *F*, or *QF* if they become quarantined). This allows us to assume that secondary infections within a household in isolation (denoted with a superscript *QS* or *Q*) play no further role in any of the transmission dynamics. As a consequence, high levels of household isolation can drive the epidemic extinct, even if within household transmission is high – an effect not achievable with the standard SEIR-type modelling approach. This improved methodology also helps to capture to some degree household depletion of susceptibles (or saturation of infection), as secondary infections in the household are incapable of generating additional household infections.

#### S1.2 Capturing social distancing

We obtained age-structured contact matrices for the United Kingdom from Prem et al. [4]. We used these contact matrices to provide information on normal levels household transmission (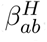, with the subscript *ab* corresponding to transmission from age group *a* against age group *b*), school-based transmission 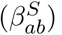, work-place transmission 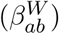 and transmission in all other locations 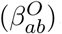.

We assume that any instigated non-pharmaceutical interventions (patterns of social-distancing or lockdown measures) leads to a reduction in the work, school and other matrices while increasing the strength of household contacts. Any given level of non-pharmaceutical interventions (NPIs), captured by the parameter *ϕ* between zero and one, therefore scales the four transmission matrices between their normal values (when *ϕ* = 0) and their value under the most severe lockdown (*ϕ* = 1).

We infer the level of NPIs as a slowly varying parameter in the MCMC processes on a weekly basis. In turn, the weekly value of *ϕ* allows us to calculate the growth rate *r* (and hence the reproductive number *R*) by an eigenvalue approach.

#### S1.3 Parameter Inference

As with any model of this complexity, there are multiple parameters that determine the dynamics. Some of these are global parameters and apply for all geographical regions, with others used to capture the regional dynamics. Some of these parameters are matched to the early outbreak data (including the resultant age-distribution of infection), however the majority are inferred by an MCMC process (Table 1).

**Table 1:**
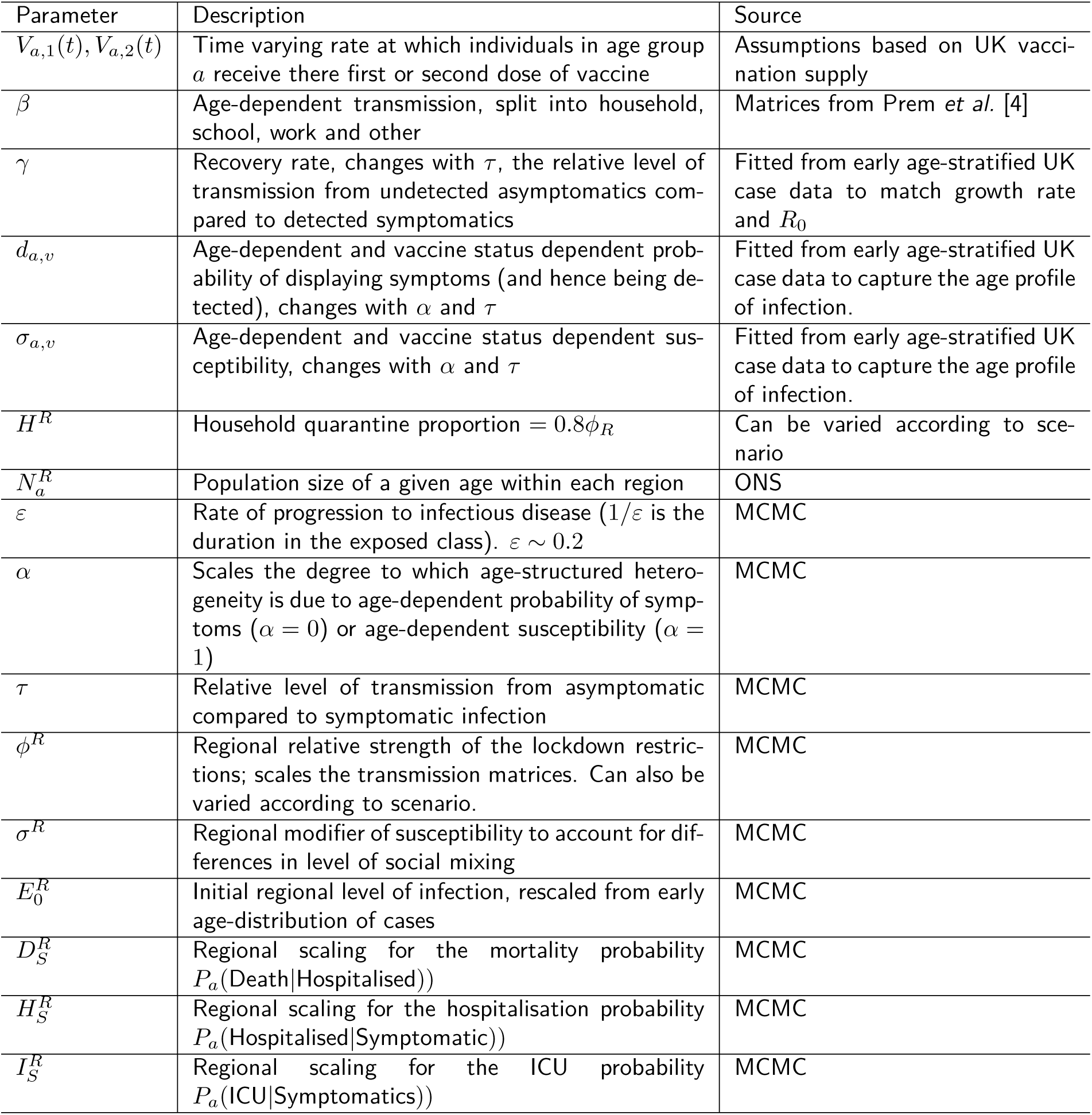
Key model parameters and their source

We would highlight that the parameters of *α* and *τ* are key in determining age-structured behaviour and are therefore essential in quantifying the role of school children in transmission [7]. We argue that a low *τ* and a low *α* are the only combination that are consistent with the growing body of data suggesting that levels of seroprevalence show only moderate variation across age-ranges [8], yet children are unlikely to display major symptoms, suggesting their role in transmission may be lower than for other respiratory infections [9, 10].

Throughout the current epidemic, there has been noticeable heterogeneity between the different regions of England and between the devolved nations. In particular, London is observed to have a large proportion of early cases and a relatively steeper decline in the subsequent lock-down than the other regions and the devolved nations. In our model this heterogeneity is captured through three regional parameters (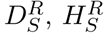 and 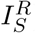) which act on the heterogeneous population pyramid of each region to generate key observables.

#### S1.4 Public Health Measurable Quantities

The main model equations focus on the epidemiological dynamics, allowing us to compute the number of symptomatic and asymptomatic infectious individuals over time. However, these quantities are not directly measured - and even the number of confirmed cases (the closest measure to symptomatic infections) is highly biased by the testing protocols at any given point in time. It is therefore necessary to convert infection estimates into quantities of interest that can be compared to data. We considered seven such quantities which we calculated from the number of new symptomatic infections on a given day 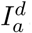.

1. **Hospital Admissions:** An age-dependent fraction of symptomatic individuals are assumed to need hospital treatment, with a distributed lag between infection and hospitalisation.
2. **ICU Admissions:** Similarly, an age-dependent fraction of symptomatic individuals are assumed to need treatment in an Intensive Care Unit. This is not a quantity that is generally reported, and therefore we cannot match our model predictions to this data source.
3. **Hospital Beds Occupied:** By convolving hospital admissions with the distributions of lengths of stay, we can estimate the number of hospital beds occupied.
4. **ICU Beds Occupied:** A similar process generates the number of occupied ICU beds.
5. **Number of Deaths:** Mortality is assumed to occur to a fraction of hospitalised individuals, with the probability of mortality dependent upon age, and occuring after a distributed lag.
6. **Proportion of Pillar 2 positives:** Given that the raw number of detected cases in any region is substantially influenced by the number of tests conducted, we consider the proportion of pillar 2 tests that are positive as a less biased figure. We assume that those symptomatically infected with COVID-19 compete with individuals suffering symptoms for other infections for the available testing capacity. This leads to proportion of pillar 2 tests that are positive being a saturating function of the number of symptomatic infections, with a single scaling parameter.

We compared these model predictions to the data by assuming that the true numbers are drawn from a negative binomial distribution with the model value as the mean, while the true proportions (Pillar 2 positives) are from a beta-binomial.

### S2 Extensions to Main Text Scenarios

In the main text we focused on a few chosen scenarios that illustrate the range of plausible behaviours, and only considered COVID-19 related deaths. Here, we show some other representative scenarios and the impact on the number of hospitalisations under all of these cases. We also display the 95% credible intervals, as defined by the variability in inferred parameters; these shaded regions contain 95% of all simulations at every point in time. (We note that any one prediction will not necessarily follow the upper or lower bound, these are envelopes that contain predictions that may wander both above and below the mean.)

#### S2.1 Alternative Step-wise NPI relaxation

We extend the graphs shown in the main paper (Figure 2(a,b)), to consider the relaxation or complete release of non-pharmaceutical interventions (NPIs). Moderate relaxation of NPIs to a level seen in early September 2020 (Figure S3(a,b)) can lead to a sustained wave of deaths. Later relaxation leads to smaller waves, as more individuals have been vaccinated, with the smallest waves occurring if there is strong infection-blocking capabilities (Figure S3(b), green line). Similarly, complete removal of NPIs in April 2021 or even January 2022 (Figure S3(c,d)) can generate extremely high waves of infection with very large numbers of deaths. Again, only late release on controls and a strong infection-blocking vaccine can mitigate the worst effects. We stress that as hospitalisations and deaths increased we would expect both national legislation and emergent behaviour to limit the spread, so these predictions should be considered as extreme worse-case scenarios.

#### S2.2 Alternative Gradual NPI-relaxation

We complement the graphs shown in the main paper (Figure 2(d,e)), with other temporal profiles of NPI relaxation; in all cases NPIs drop steady from their value estimated in mid-January 2021 to zero. Figure S4(e) summarises the combined impact of relaxation and infection efficacy. For a given set of vaccine characteristics, we obtained the greatest benefit from a long slow release of NPIs (Figure S4(c)), whereas a rapid early release (Figure S4(a)) is predicted to cause the most deaths. In general there are minimal differences between vaccines with low infection efficacy (0% compared to 35%), and it is only for the highest infection efficacy of 85% that deaths are substantially reduced.

#### S2.3 Impact of Vaccination and NPI-release on Hospital Admissions

Corresponding figures to Figure S3 and S4 are given for the number of predicted hospital admissions. These show a very similar pattern to deaths.

**Figure S3:**
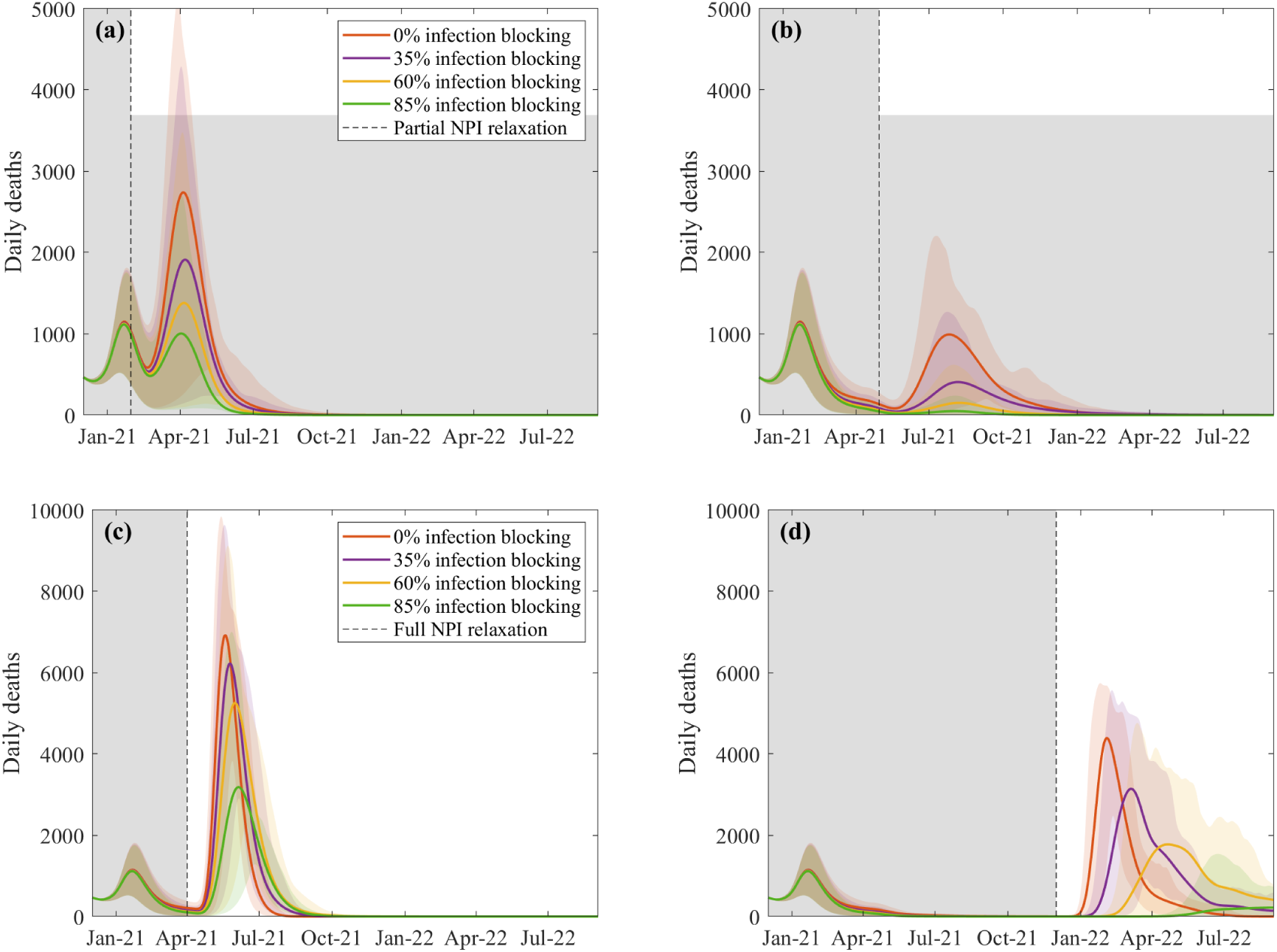
Predicted daily deaths in the UK following the start of an immunisation program and relaxation or removal of NPIs. Panels **(a)** and **(b)** show the effect of partial NPI measures down to those seen in early September 2020 (generating *R* ∼ 1.2 − 1.6 dependent on the region and level of the new variant) from January or April 2021 respectively, while panels **(c)** and **(d)** show the complete removal of all NPI measures (leading to *R* ∼ 2.4 − 3.4) from either April or at the final stages of vaccination in July 2021. Shaded regions show the 95% credible intervals.

**Figure S4:**
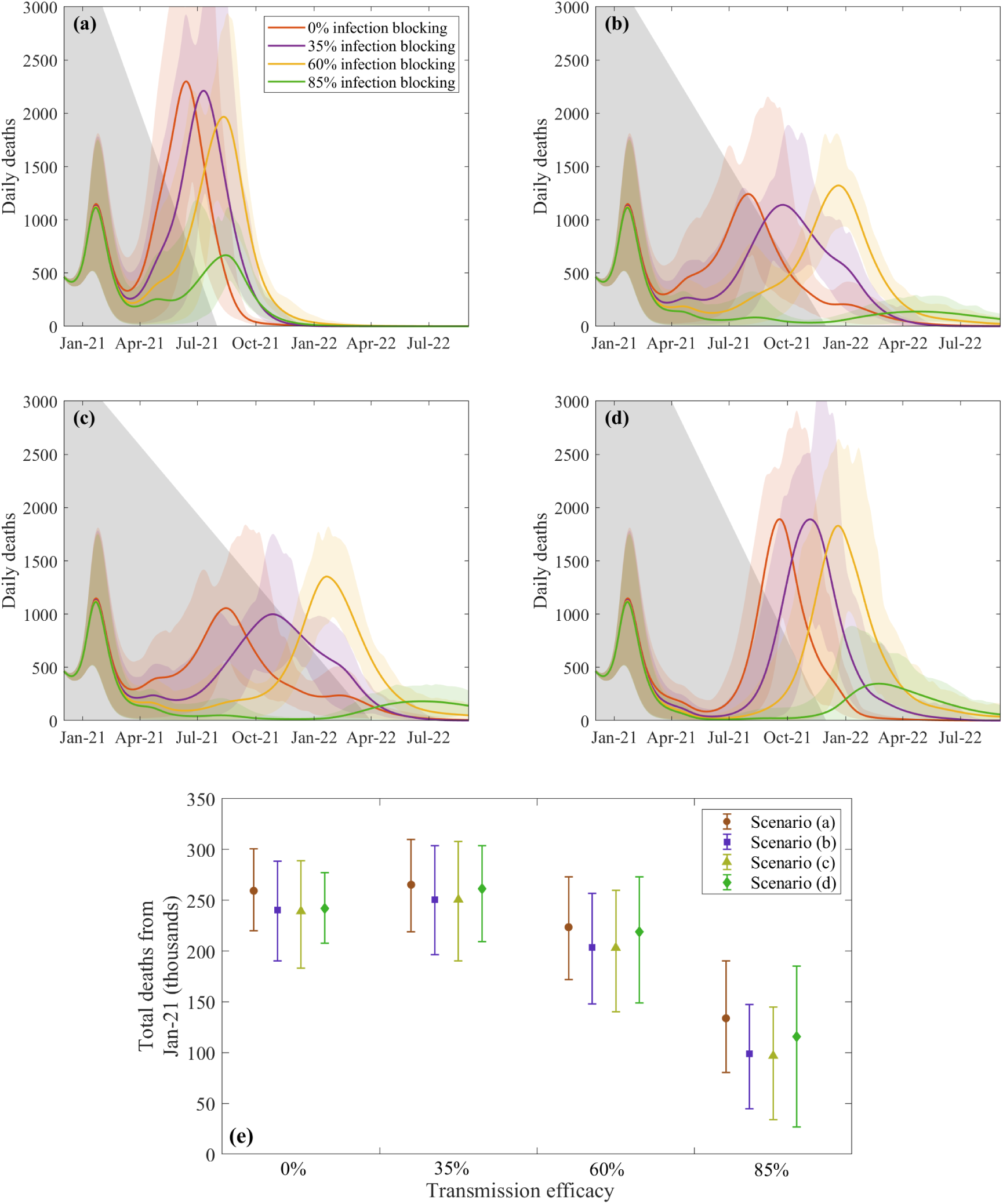
Effect of gradual relaxation of NPI measures on deaths across the UK following the start of vaccination. In panels **(a)-(d)** different relaxation scenarios are shown with NPIs reduced linearly from December levels down to complete release over different time periods represented by the height of the grey shading. Panel **(e)** compares the total predicted deaths from Jan-21 onwards between the scenarios for each vaccine efficacy.

**Figure S5:**
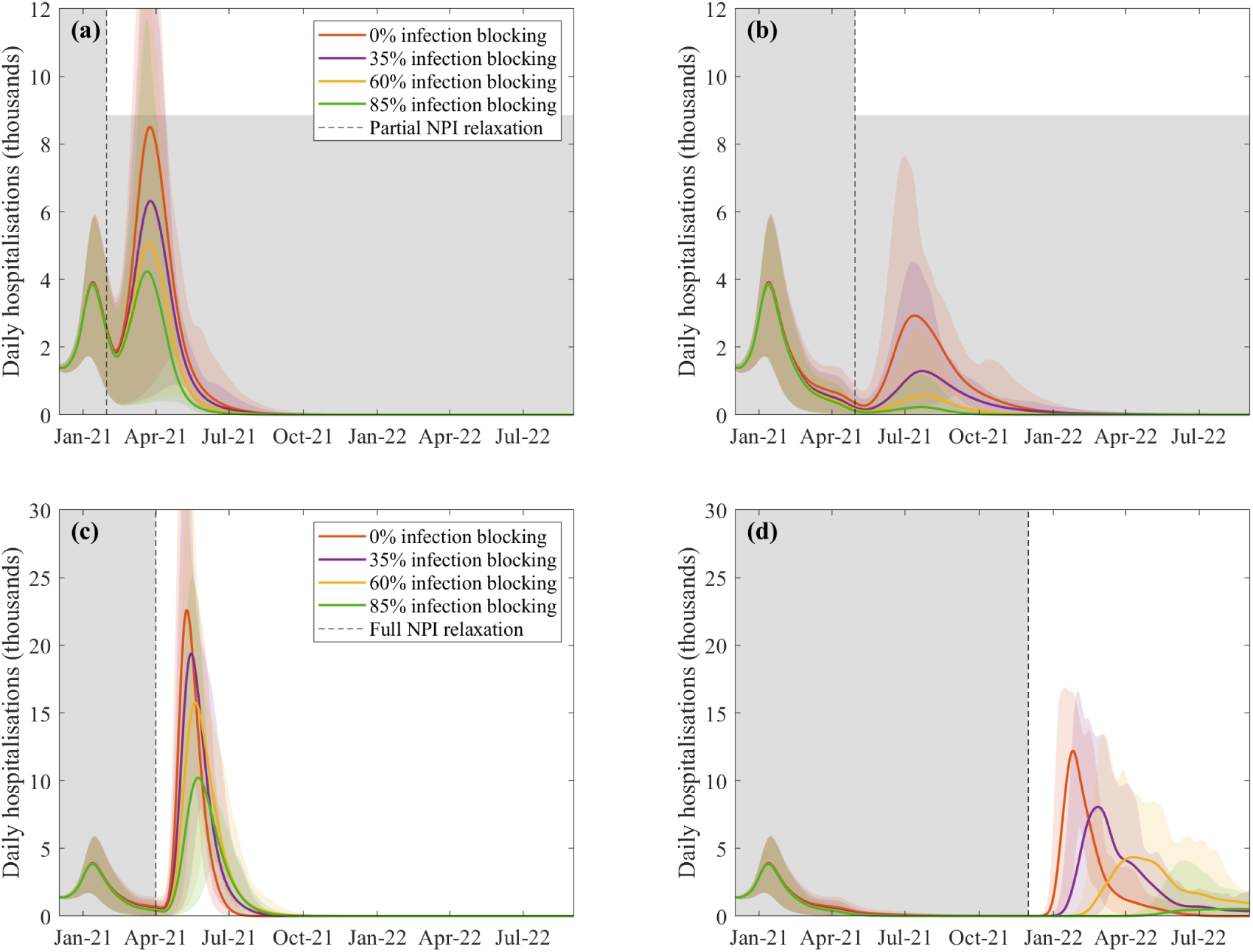
Predicted daily hospitalisations in the UK following the start of an immunisation program and relaxation or removal of NPIs. Panels **(a)** and **(b)** show the effect of partial NPI measures down to those seen in early September 2020 (generating *R*∼ 1.2 − 1.6 dependent on the region and level of the new variant) from January or April 2021 respectively, while panels **(c)** and **(d)** show the complete removal of all NPI measures (leading to *R* ∼ 2.4 − 3.4) from either April or at the final stages of vaccination in July 2021.

**Figure S6:**
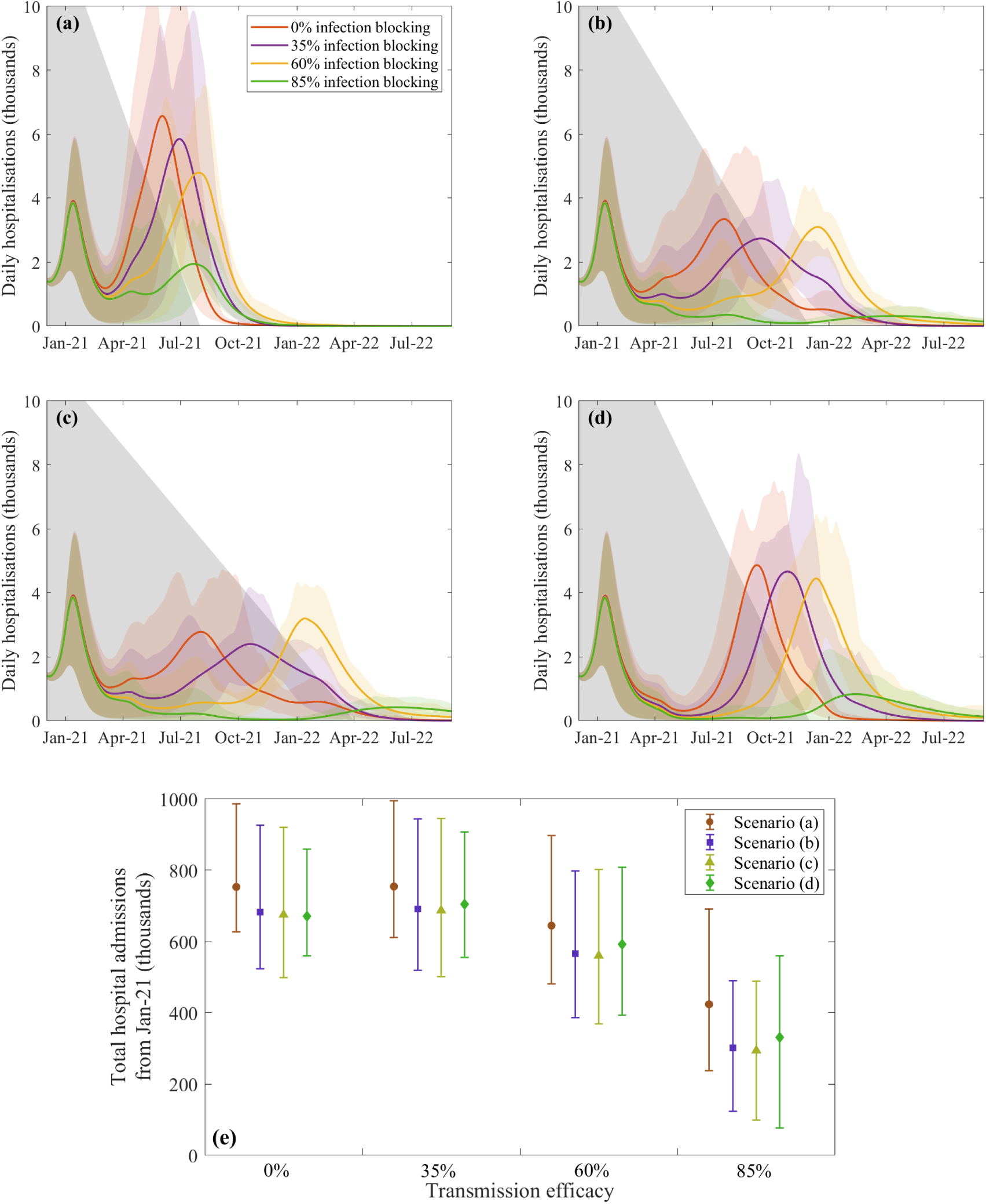
Effect of gradual relaxation of NPI measures on hospital admissions across the UK following the start of vaccination. In panels **(a)-(d)** different relaxation scenarios are shown with NPIs reduced linearly from December levels down to complete release over different time periods represented by the height of the grey shading. Panel **(e)** compares the total predicted hospital admissions from Jan-21 onwards between the scenarios for each vaccine efficacy.

